# Bacterial Pneumonia and Respiratory Culture Utilization among Hospitalized Patients with and without COVID-19 in New York City

**DOI:** 10.1101/2022.02.08.22270591

**Authors:** Maxwell D. Weidmann, Gregory J. Berry, Jason E. Zucker, Simian Huang, Magdalena E. Sobieszczyk, Daniel A. Green

**Affiliations:** Department of Pathology & Cell Biology, Columbia University Irving Medical Center, New York, NY; Division of Infectious Diseases, Department of Internal Medicine, Columbia University Irving Medical Center, New York, NY

**Keywords:** COVID-19, SARS-CoV-2, Pneumonia, Co-infection, Respiratory Culture

## Abstract

COVID-19 is associated with prolonged hospitalization and a high risk of intubation, which raises concern for bacterial co-infection and antimicrobial resistance. Previous research has shown a wide range of bacterial pneumonia rates for COVID-19 patients in a variety of clinical and demographic settings, but none have compared hospitalized COVID-19 patients to patients testing negative for SARS-CoV-2 in similar care settings. We performed a retrospective cohort study on hospitalized patients with COVID-19 testing from 10 March 2020 to 31 December 2020. A total of 19,219 patients were included, of which 3,796 tested positive for SARS-CoV-2. We found a 2.6-fold increase (p < 0.001) in respiratory culture ordering in COVID-19 patients. On a per-patient basis, COVID-19 patients were 1.5-fold more likely than non-COVID patients to have abnormal respiratory cultures (46.8% vs. 30.9%, p < 0.001), which was primarily driven by patients requiring intubation. Among patients with pneumonia, a significantly higher proportion of COVID-19 patients had ventilator-associated pneumonia (VAP) relative to non-COVID patients (85.7% vs 55.1%, p <0.001), but a lower proportion had community-acquired (12.2% vs 22.1%, p < 0.01) or hospital-acquired pneumonia (2.1% vs. 22.8%, p < 0.001). There was also a significantly higher proportion of respiratory cultures positive for MRSA, *K. pneumoniae*, and antibiotic-resistant organisms in COVID-19 patients. Increased rates of respiratory culture ordering for COVID-19 patients therefore appear to be clinically justified for patients requiring intubation, but further research is needed to understand how SARS-CoV-2 increases the risk of VAP.

## Introduction

Concern for bacterial co-infection among COVID-19 patients resulted in empirical antimicrobial therapy given to a large proportion of patients hospitalized at our medical center during 2020 [1]. While a relatively high rate of bacterial and influenza co-infection has characterized hospitalized influenza patients, with estimates ranging from 11% to 35% [2, 3], less is known about the rate of bacterial pneumonia in hospitalized COVID-19 patients. Further, in influenza, bacterial co-infection has been found to be more frequently of community-origin, with one study of hospitalized adults in the US finding that more than 54% of co-infections in adults were diagnosed within the first 48 hours of hospitalization [3]. Several studies have examined bacterial co-infection for COVID-19 patients in inpatient and ICU settings, generally finding lower rates than for influenza patients [4-7]. Of the two largest such studies, one found very low rates of community-acquired pneumonia (CAP) (1.5%) [7], while the other found an overall bacterial pneumonia rate of 2.1%, with the overwhelming majority being HAP or VAP [6, 8]. One study among ICU patients found much higher rates of patients with abnormal respiratory cultures (28%), however 90% of these patients required mechanical ventilation [9].

Therefore, when all hospitalized patients are considered together, rates of bacterial pneumonia appear lower in COVID-19 patients relative to hospitalized influenza patients, and in contrast to influenza, few of these infections are community-acquired. However, none of these studies have assessed the concurrent rate of bacterial co-infections, including HAP and VAP rates, in patients testing negative for SARS-CoV-2 during the same period. This lack of a direct comparison between COVID-19 and non-COVID-19 infected patients makes it difficult to determine whether the highly disparate reported rates of bacterial co-infection represent the heterogeneity of the patient populations studied, or a truly elevated risk of bacterial pneumonia associated with SARS-CoV-2 infection.

Additionally, respiratory culture utilization among COVID-19 patients has not been previously studied. Understanding the rates of respiratory culture ordering and positivity among COVID-19 patients is needed to determine whether culture ordering is appropriate among hospitalized COVID-19 patients, and to understand the risk of HAP and VAP in this population. To study this further, we compared rates of bacterial pneumonia, respiratory culture ordering, bacterial etiologies, and antibiotic resistance between patients testing positive and negative for COVID-19 in the same hospital setting.

## Materials and Methods

A retrospective cohort study was conducted on all hospitalized patients with SARS-CoV-2 testing performed at Columbia University Irving Medical Center (CUIMC) located in New York City from 10 March 2020 to 31 December 2020. SARS-CoV-2 RT-PCR testing was performed in-house using the following assays: Cobas SARS-CoV-2 (Roche Molecular Systems, Inc., Branchburg, NJ), Xpert Xpress SARS-CoV-2 (Cepheid, Sunnyvale, CA), and a laboratory-developed test from the Wadsworth Center at the New York State Department of Health.

Data were extracted from medical records to compare respiratory culture utilization and culture results between COVID-19 and non-COVID patients; these data included time of SARS-CoV-2 testing, SARS-CoV-2 test results, dates of admission and discharge, intubation status, number of respiratory cultures performed, time of respiratory culture orders, and results of respiratory culture testing, including time of culture results, bacterial species recovered, and antimicrobial susceptibility.

Rates of respiratory culture utilization and culture positivity were compared between COVID-19 and non-COVID patients, as was the distribution of recovered species, antimicrobial susceptibility profiles, intubation status and ordering time of positive cultures during hospital admission to stratify pneumonia cases into community-acquired (<= 2 days) vs. hospital-acquired (> 2 days) vs ventilator-associated (>2 days after intubation) [8]. A positive respiratory culture was defined as any potential respiratory pathogen recovered from culture, whereas a negative result was defined as either no growth, when cultures were resulted as “mixed commensal microbiota,” or when only yeast were isolated. Respiratory cultures were excluded if SARS-CoV-2 testing was performed after respiratory cultures were ordered to ensure that all culture results analyzed were from patients of known COVID-19 status at time of testing.

Chi-squared testing was used to identify statistically significant differences in rates of respiratory culture ordering, positivity, antibiotic resistance, and CAP, HAP or VAP in COVID-19 positive and negative patients. A two-tailed Student’s t-test was used to assess statistical significance in time to respiratory culture positivity (when results of continuous variables were being compared). Data analysis was initially performed using Microsoft Excel, with statistical tests performed using R and R-Studio for reproducibility with scripts included in supplemental data.

## Results

### Demographics of SARS-CoV-2 positive and negative populations

A total of 19,275 participants were initially included; 3,805 (19.7%) tested positive for SARS-CoV-2 and 15,470 (80.3%) tested negative. 56 patients were excluded due to respiratory cultures ordered prior to SARS-CoV-2 testing, leaving 19,219 participants for study analysis. SARS-CoV-2 positivity rates varied widely over the study period, from a peak of nearly 60% in April 2020 to a low of just over 3% in September 2020 (**Supplemental Figure 1)**. Demographic characteristics of included patients are shown in **Table 1**. Compared to non-COVID patients, COVID-19 patients were older (p < 0.001), more likely to be male (p < 0.001), more likely to be Hispanic/Latino, less likely to be white, and were more likely to receive care in the ICU (p < 0.001). The mortality rate was also 5.6-fold higher (16.9% vs. 3.0%, p < 0.001) and intubation rate 2.9-fold higher (16.6 vs. 5.7, p < 0.001) for COVID-19 patients.

**Table 1:**
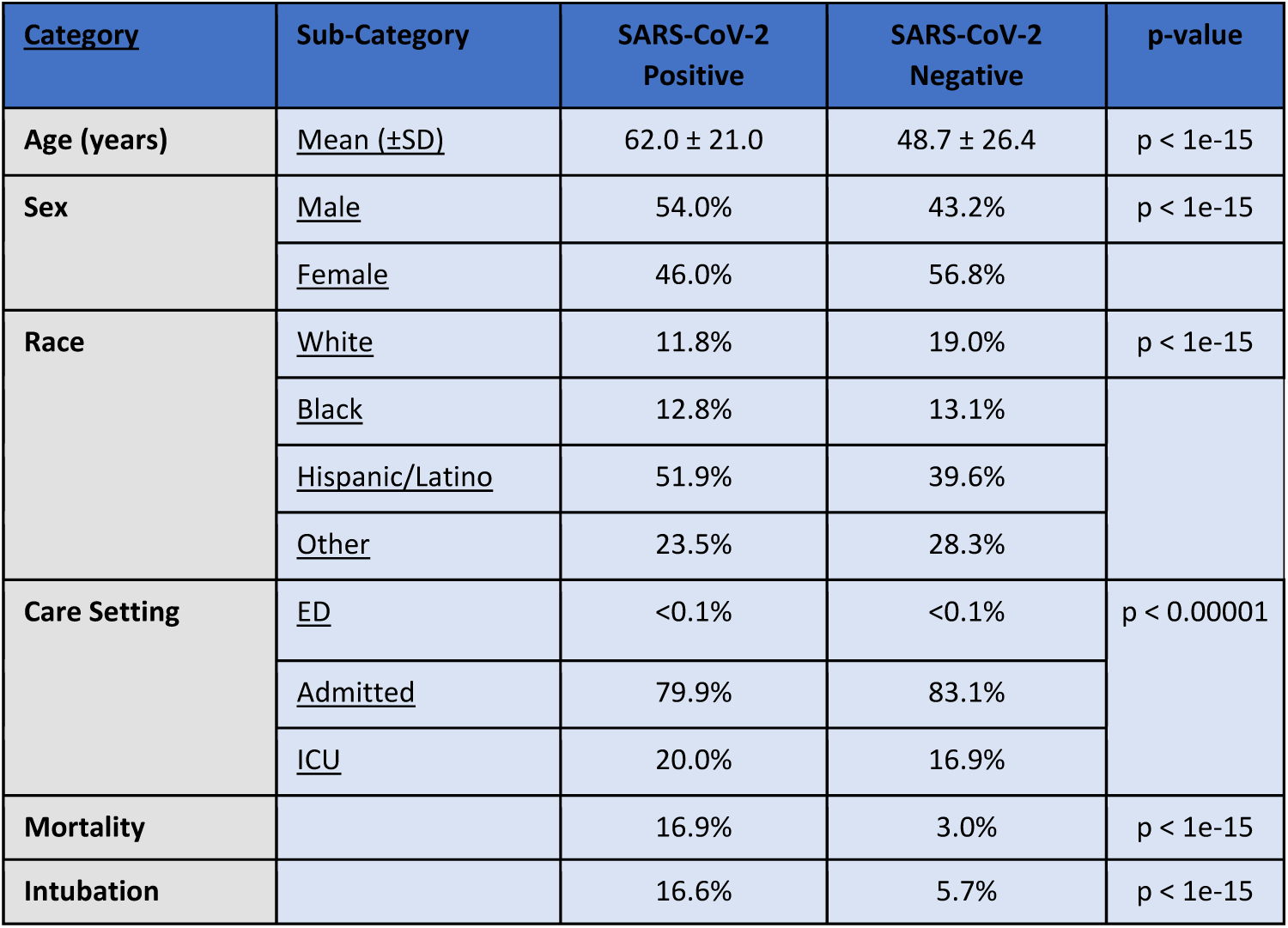
Demographics of SARS-CoV-2 Positive and Negative Patient Population.

**Figure 1:**
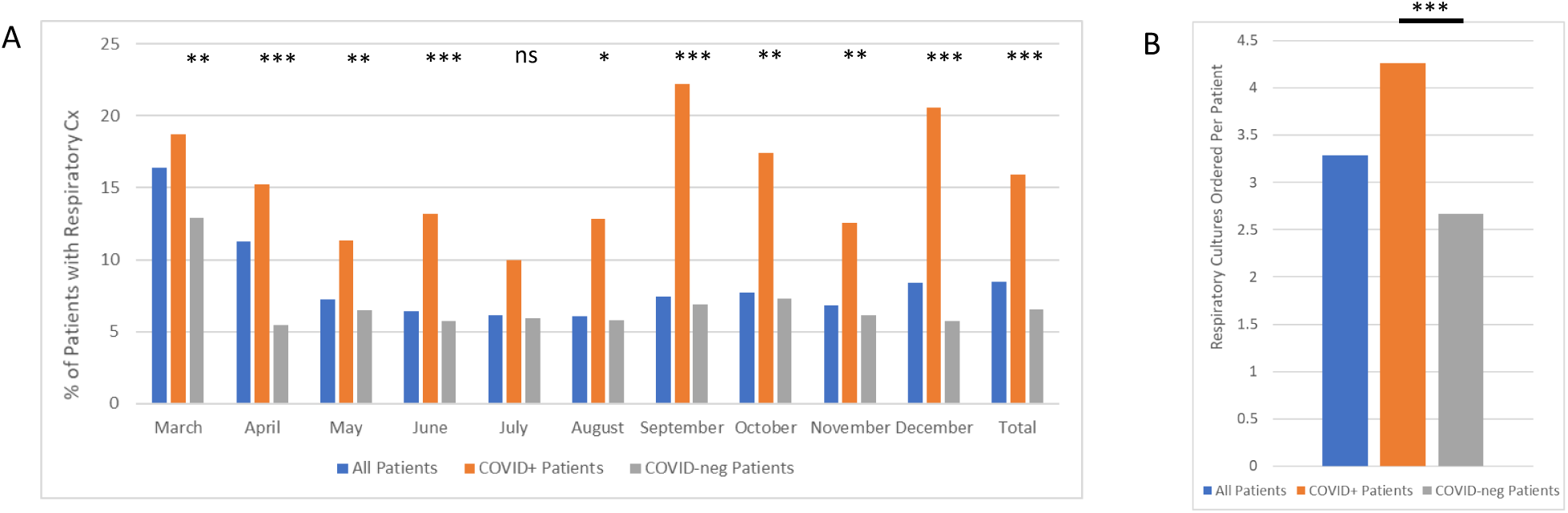
Respiratory culture ordering by COVID-19 status. A) Percentage of patients with respiratory cultures ordered, by month and total. B) Average number of respiratory cultures ordered per patient. * p < 0.05, ** p < 0.01, *** p < 0.001, ns = not significant.

### Increased rates of respiratory culture ordering in COVID-19 patients

COVID-19 patients were 2.6 times as likely to have respiratory cultures ordered compared to non-COVID patients (16.0 % vs 6.2%, p < 0.001, **Table 2**). Although the rate of respiratory culture ordering varied by month (**Figure 1A**), significantly higher ordering was seen among COVID-19 patients across all months except July 2020. Increased ordering was also reflected in a higher number of cultures ordered per patient for COVID-19 patients relative to non-COVID patients (4.3 vs 2.7 per patient, p <0.001, **Figure 1B**).

**Table 2:** Respiratory Culture Ordering.

| <u>Metric</u> | **Total** | **SARS-CoV-2 Positive** | **SARS-CoV-2 Negative** | **p-value** |
| --- | --- | --- | --- | --- |
| **Total Patients** | 19219 | 3796 | 15423 |  |
| **Total cultures ordered** | 5152 | 2587 | 2565 |  |
| **Patients with respiratory cultures ordered** | 1569 (8.2%) | 607 (16.0%) | 962 (6.2%) | p < 1e-15 |
| **Cultures ordered per patient** | 3.3 | 4.3 | 2.7 | p < 1e-14 |
| **Intubated patients with respiratory cultures ordered** | 867 (57.5%) | 451 (71.5%) | 416 (47.4%) | p < 1e-15 |
| **Non-intubated patients with respiratory cultures ordered** | 792 (4.4%) | 195 (6.1%) | 597 (4.1%) | p < 1e-5 |

A much higher rate of respiratory culture ordering was seen overall among intubated patients (57.5%) vs non-intubated patients (4.4%). Among intubated patients, however, ordering was still significantly higher for those with COVID-19 compared to non-COVID patients (71.5% vs. 47.4%, p < 0.001, **Table 2, Supplementary Figure 2**). In addition, among patients who did not require intubation, there was also a significantly higher rate of respiratory culture ordering for COVID-19 relative to non-COVID patients (6.1 vs 4.1%, p < 0.001, **Supplementary Figure 2D & Table 2**).

### Higher likelihood of respiratory culture positivity among intubated COVID-19 patients

Overall, 7.6% of COVID-19 patients had positive respiratory cultures vs. 1.8% for non-COVID patients (**Figure 2, Table 3**). Among patients who had respiratory cultures ordered, COVID-19 patients were also more likely than non-COVID patients to have an abnormal culture (46.8% vs 30.9%, p < 0.001). Even accounting for the higher rate of respiratory culture ordering among COVID-19 patients, on a per culture basis, COVID-19 patients still had a higher percentage of positive cultures (33.4% vs 26.9%, p < 0.001, **Table 3**).

**Table 3:** Respiratory Culture Positivity.

| **<u>Metric</u>** | **Total** | **SARS-CoV-2**<br>**Positive** | **SARS-CoV-2**<br>**Negative** | **p-value** |
| --- | --- | --- | --- | --- |
| **Total Patients** | 19219 | 3796 | 15423 |  |
| **Intubated Patients** | 1509<br>(7.9%) | 631<br>(16.6%) | 878<br>(5.7%) |  |
| **Patients who had abnormal**<br>**respiratory cultures** | 572 (3.0%) | 287 (7.6%) | 285 (1.8%) | p < 1e-15 |
| **% of patients with abnormal**<br>**cultures among patients with**<br>**cultures ordered** | 37.0% | 46.8% | 30.9% | p < 1e-9 |
| **Cultures that resulted as abnormal** | 1554<br>(30.2%) | 863 (33.4%) | 691 (26.9%) | p < 1e-6 |
| **Intubated patients with Ventilator-**<br>**associated pneumonia (VAP) (%)** | 403 (26.7%) | 246 (39.0%) | 157 (17.9%) | p < 1e-15 |
| **% of intubated patients with**<br>**respiratory cultures ordered and**<br>**Ventilator-associated pneumonia**<br>**(VAP)** | 57.4% | 62.0% | 51.5% | p < 0.01 |
| **Non-intubated patients who had**<br>**abnormal respiratory cultures (%)** | 133 (0.7%) | 24 (0.7%) | 109 (0.7%) | p = 1 |
| **% of non-intubated patients with**<br>**respiratory cultures ordered that**<br>**resulted as abnormal** | 16.8% | 12.3% | 18.3% | p = 0.069 |

**Figure 2.**
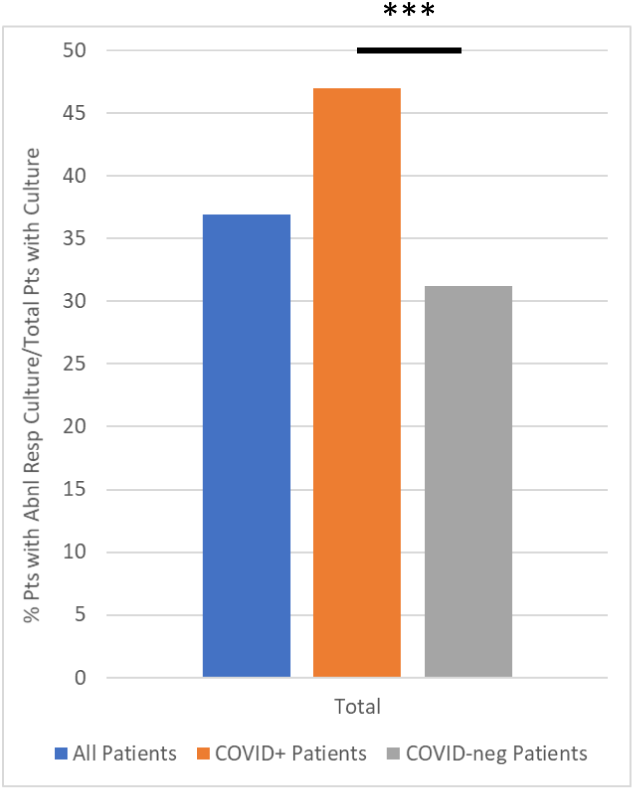
Abnormal respiratory culture rates by COVID-19 status. Percentage of patients with abnormal respiratory cultures of those with any respiratory culture ordered. *** p < 0.001

The higher rate of abnormal cultures among COVID-19 patients was primarily driven by patients requiring intubation. A total of 26.7% of all intubated patients had abnormal respiratory cultures collected more than 48 hours after intubation, qualifying as ventilator-associated pneumonia (VAP). Amongst intubated patients, VAP was more than twice as likely in COVID-19 patients relative to non-COVID patients (39.0% vs. 17.9%, p < 0.001, **Supplementary Figure 2E, Table 3**). When looking at only intubated patients who had respiratory cultures ordered, there remained a significantly higher proportion of COVID-19 positive patients with VAP compared to non-COVID patients (62.0% vs. 51.5%, p < 0.01, **Supplementary Figure 2E, Table 3**) However, among non-intubated patients, there were no significant differences by COVID-19 status in the proportion of patients with respiratory cultures ordered that resulted as abnormal or the proportion of total patients with abnormal respiratory culture results (**Supplementary Figure 2E-F, Table 3)**.

Among culture-positive patients, COVID-19 patients had a significantly lower rate of CAP, defined as abnormal cultures within the first two days of admission, compared to non-COVID patients (12.2% vs 22.1%, p < 0.01, **Figure 3, Table 4**). Conversely a significantly higher proportion of pneumonia cases also occurred at least two days after admission for COVID-19 positive patients relative to non-COVID patients (**Figure 3**), and this difference was particularly pronounced in patients with positive cultures at least 10 days after admission (64.8% vs. 42.1%, p < 0.001). When we looked specifically at HAP, defined as abnormal respiratory cultures that resulted more than two days after admission but not in patients who had been ventilated for more than two days, we found a significantly lower rate among COVID-19 patients relative to non-COVID patients (2.1% vs. 22.8%, p < 0.001, **Figure 3, Table 4**). The overwhelming majority (85.7%) of bacterial pneumonia cases among COVID-19 patients were ventilator-associated, and this was significantly higher than the rate of VAP in non-COVID patients with bacterial pneumonia (55.1%, p < 0.001, **Figure 3, Table 4**).

**Table 4:**
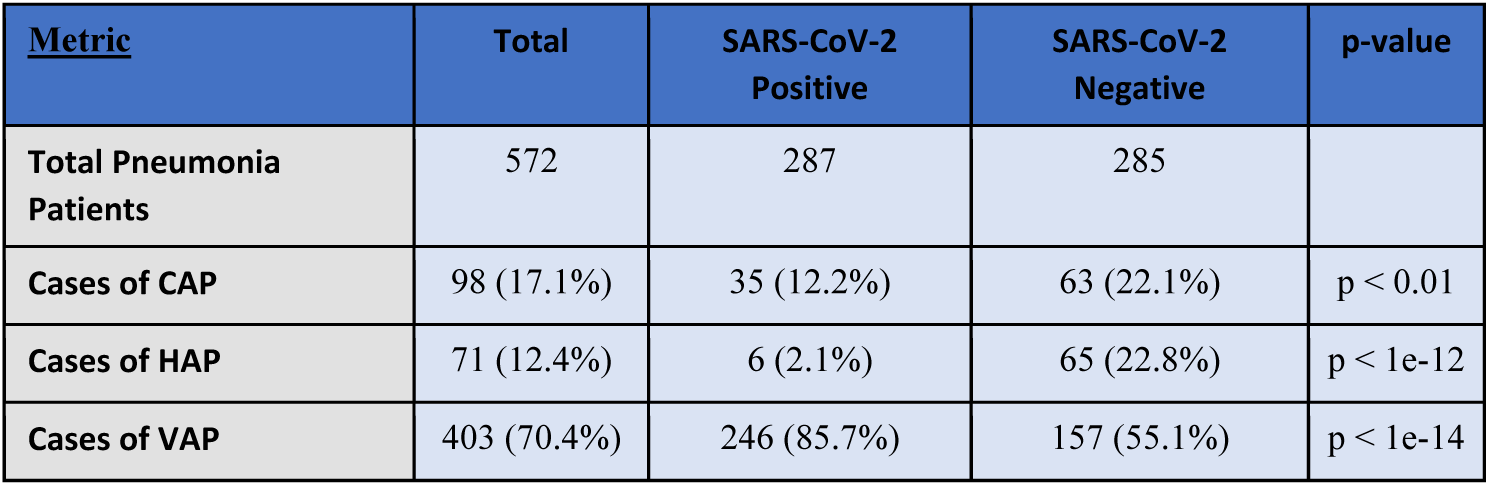
Bacterial Pneumonia Categories.

| **<u>Metric</u>** | **Total** | **SARS-CoV-2**<br>**Positive** | **SARS-CoV-2**<br>**Negative** | **p-value** |
| --- | --- | --- | --- | --- |
| **Total Pneumonia**<br>**Patients** | 572 | 287 | 285 |  |
| **Cases of CAP** | 98 (17.1%) | 35 (12.2%) | 63 (22.1%) | p < 0.01 |
| **Cases of HAP** | 71 (12.4%) | 6 (2.1%) | 65 (22.8%) | p < 1e-12 |
| **Cases of VAP** | 403 (70.4%) | 246 (85.7%) | 157 (55.1%) | p < 1e-14 |

**Figure 3:**
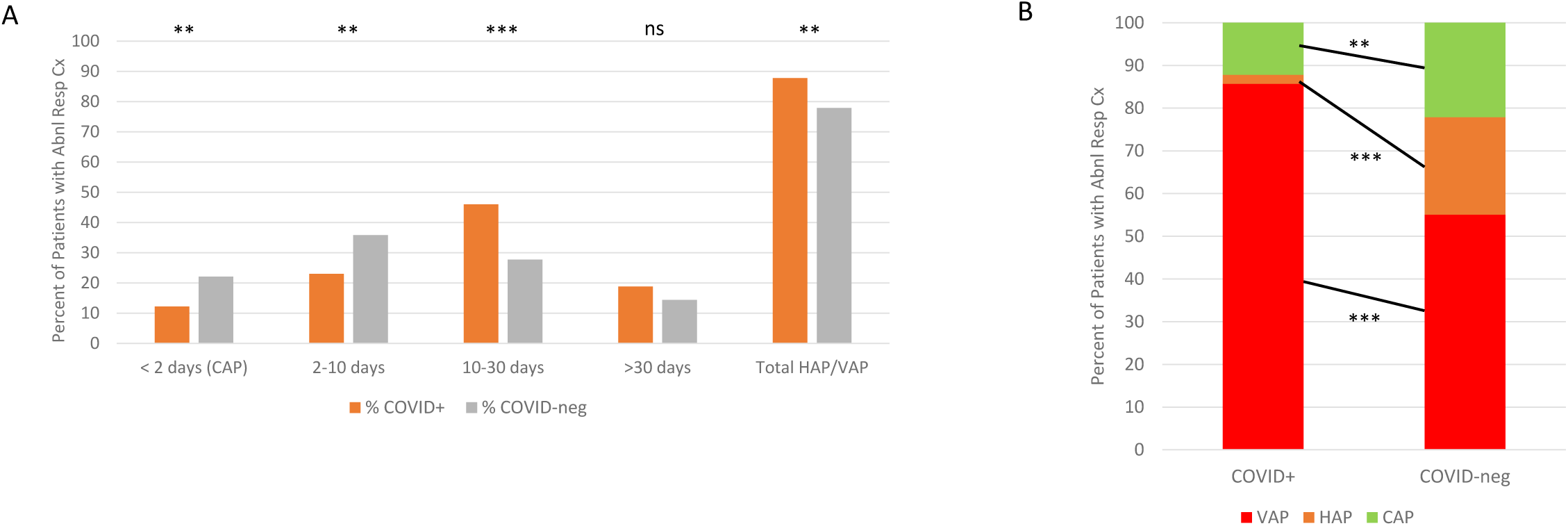
Time from admission to respiratory culture positivity. A) Percentage of total COVID-19 or non-COVID patients with abnormal respiratory culture grouped by time interval from admission to respiratory culture ordering comparing community acquired pneumonia (CAP, <48hrs) to hospital-acquired (HAP) or ventilator-associated pneumonia (VAP). B) Percentage of total COVID-19 or non-COVID patients with abnormal respiratory culture who fit criteria for CAP, HAP or VAP [8]. * p < 0.05, ** p < 0.01, *** p < 0.001

### Bacterial etiologies of positive respiratory cultures

Respiratory cultures grew a wide range of bacterial pathogens in our patient population (**Supplementary Table 1**). The most common organism overall was *P. aeruginosa* (**Figure 4**). For COVID-19 patients, the most common organisms in descending order were *S. aureus, P. aeruginosa* and *K. pneumonia*. For non-COVID patients, the most common organisms in descending order were *P. aeruginosa, S. aureus* and *K. Pneumoniae* (**Figure 4**). Among patients with abnormal respiratory cultures, methicillin-resistant *S. aureus* infections (MRSA) were more common for COVID-19 patients (8.9% vs. 5.4%, p < 0.05, **Figure 4**), and there was a significant increase in the proportion of overall COVID-19 positive vs. negative patients with MRSA respiratory infections (1.05% vs. 0.15%, p < 0.001). There was also a significantly higher proportion of COVID-19 patients whose cultures grew *K. pneumoniae* (26.1% vs. 16.3%, p < 0.01, **Figure 4)**.

**Figure 4:**
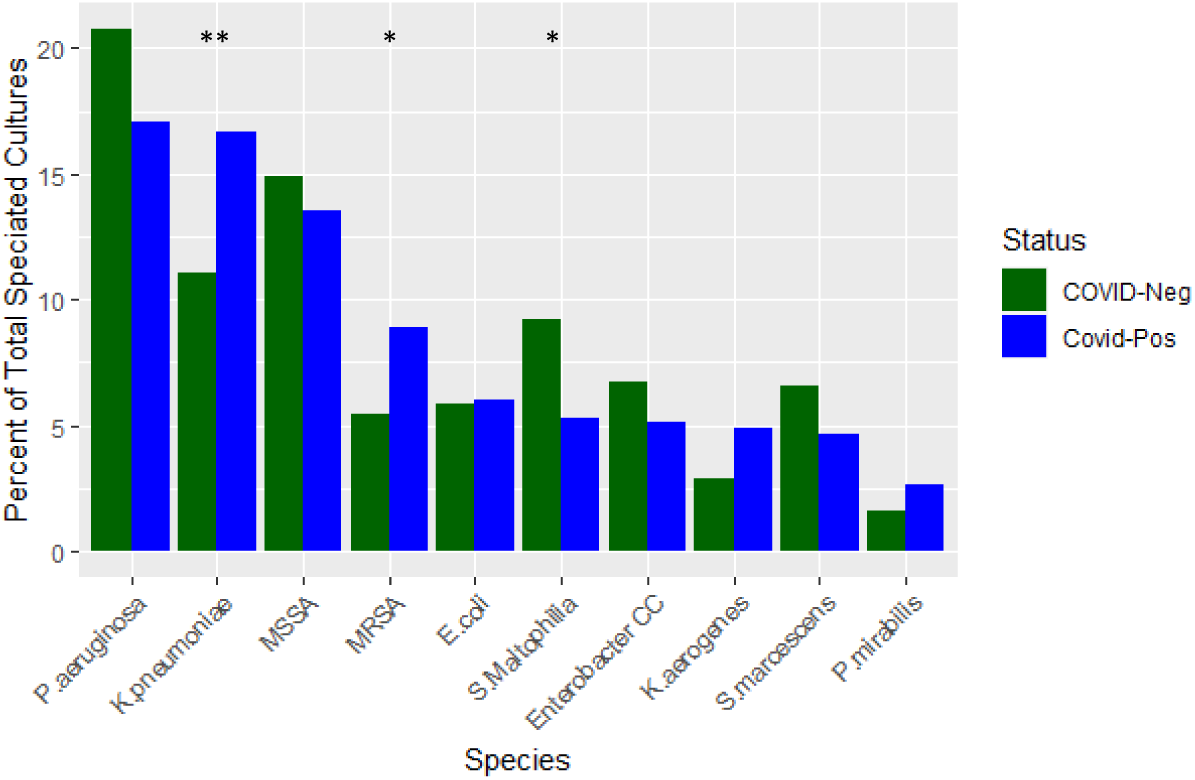
Bacterial species identified in abnormal respiratory cultures in COVID-19 positive vs. negative patients. Percent of total patients in each category of COVID-19 status with the 10 most common species for COVID-19 patients in descending order. * p < 0.05, ** p < 0.01, *** p < 0.001

### Antibiotic resistance in COVID-19 bacterial pneumonia

In agreement with our finding of increased rates of MRSA-positive respiratory cultures in COVID-19 patients, we found increased rates of resistance to penicillin class antibiotics (penicillin, ampicillin, oxacillin), 63.4 vs. 50.1% of COVID-19 positive vs. negative patients who had an abnormal culture result (p < 0.05, **Supplemental Figure 3**). Rates of penicillin-class resistance amongst gram-positive organisms were higher amongst COVID-19 patients (51.8 vs. 40.7%), but this difference was not significant. Relative to non-COVID patients, COVID-19 patients showed no significant increases in rates of Enterbacterales resistant to 3^rd^-generation cephalosporins, carbapenem-resistant Enterobacterales, and carbapenem-resistant *P. aeruginosa or A. baumannii*.

## Discussion

During the first 10 months of the COVID-19 pandemic we found significantly higher utilization of respiratory cultures for COVID-19 patients compared to non-COVID patients at our NYC medical center. Yet despite the increased ordering, intubated COVID-19 patients still had a higher percentage of abnormal respiratory cultures than intubated non-COVID patients, suggesting that COVID-19 patients have an elevated risk for VAP. Bacterial pneumonia in COVID-19 patients was much more likely to be ventilator-associated and less likely to be community acquired when compared to non-COVID patients. When excluding intubated patients, rates of hospital-acquired pneumonia were lower among COVID-19 relative to non-COVID patients. This study is the first to directly assess respiratory culture ordering and positivity in COVID-19 and non-COVID patients in the same hospital. Given the higher rate of positivity amongst intubated COVID-19 patients, respiratory culture ordering appears warranted for this subset of hospitalized COVID-19 patients in whom there is a higher suspicion for bacterial pneumonia.

While more than 80% of respiratory co-infections in COVID-19 patients occurred more than two days into hospitalization, and the majority of these occurred well into hospitalization (> 10 days), nearly all these cases qualified as VAP and were therefore attributable to intubation rather than other sources of hospital-acquired infection. Only 0.7% of non-intubated COVID-19 patients were found to have bacterial pneumonia, and when this was normalized to respiratory culture ordering, the rate was lower than that for non-intubated non-COVID patients. We also found similarly low rates of CAP in COVID-19 patients compared to what has previously been cited in the literature. A recent study of hospitalized COVID-19 patients looking specifically at rates of bacterial co-infection within the first three days of hospitalization and finding 1.1% with probable infection, and additional 12.4% with possible co-infection, when all types of infection were considered [10]. Looking specifically at respiratory bacterial co-infections, our study found 1.2% of COVID-19 patients with positive cultures in the first three days of hospitalization, in contrast to 6.3% of COVID-19 patients with abnormal cultures representing HAP or VAP by this definition. The low rate of community-acquired respiratory infections we found here further supports these authors’ conclusions that empiric antibiotic therapy for COVID-19 patients is generally not indicated during early hospitalization [10, 11]. Similarly, our finding that non-intubated patients had lower rates of bacterial pneumonia, when those rates were normalized by respiratory culture ordering, argues against routine respiratory culture and empiric pneumonia therapy in these patients. However, the relatively high rates of VAP that we found in COVID-19 patients supports a low threshold for respiratory culture ordering and initiation of antimicrobial therapy for intubated patients.

Abnormal respiratory cultures of COVID-19 patients were enriched for pathogens associated with hospital-acquired infection with a generally similar proportion non-COVID patients (**Figure 4**). However, there were significant increases in both *Klebsiella pneumoniae* and MRSA respiratory infections, which we interpreted as further corroborating an increased level of hospital-acquired infections for COVID-19 positive patients, particularly relative to the total number of patients testing positive. Overall rates of resistance to penicillin-class antibiotics were significantly higher in respiratory isolates from COVID-19 positive patients, which corroborates data from other studies demonstrating high rates of MRSA and MDR bacterial infections in COVID-19 patients [6, 12]. One such study also showed that there were significant increases in antibiotic resistance by the duration of hospital stay, including rates of MRSA, vancomycin resistant Enterococcus (VRE), ceftriaxone-resistant Enterbacterales, carbapenem-resistant Enterobacterales and carbapenem-resistant *P. aeruginosa or A. baumannii* [12]. Interestingly, we did not find significant increase in resistance rates between COVID-19 positive and negative patients for ceftriaxone-resistant Enterbacterales, carbapenem-resistant Enterobacterales and carbapenem-resistant *P. aeruginosa or A. baumannii*. However, further study is necessary to understand the mechanism by which MRSA rates are increased by SARS-CoV-2 infection, and whether this is via direct effect of the virus on the host environment or indirectly through affecting host exposure to drug resistant pathogens.

Some limitations of this study include its representation of data from a single institution only. There were also significant baseline demographic differences in the COVID-19 positive and negative populations. Inadequate SARS-CoV-2 testing capacity also makes results from March of 2020 difficult to interpret, as false negative rates may have been increased in this setting. Prior to March 23^rd^, 2020, not all patients admitted to CUIMC were tested for SARS-CoV-2 due to limited capacity, and this population of unknown COVID-19 status was not included in our data. The data available did not include sensitivities for several antibiotic classes, such as fluoroquinolones, aminoglycosides and macrolides, that would have been helpful in a more comprehensive characterization of resistance in our population of patients with bacterial pneumonia. In addition, the data analyzed here represents one period early in the pandemic, during which time rates of COVID-19 varied greatly in the population (Supplemental Figure 1). The COVID-19 patient population represented here is largely composed of those infected during the initial surge of infections in NYC from March-May of 2020 and may not represent later stages of the pandemic during which different SARS-CoV-2 variants have become dominant.

In summary, the data presented here suggests that COVID-19 infection resulted in higher rates of bacterial co-infection relative to patients who tested negative during the same period, but that these infections were seen in the setting of prolonged hospitalization and intubation, rather than increased rates of CAP as seen with other viruses such as influenza. Possible explanations for these findings include longer periods of hospitalization for COVID-19 patients, inadequate infection control practices during acute phases of the pandemic, or direct effects of SARS-CoV-2 itself on the pathogenesis of bacterial pneumonia during intubation. Based on these findings, respiratory culture ordering is indicated for COVID-19 patients with prolonged hospitalization and ventilator dependence. However, further study is necessary to understand why SARS-CoV-2 predisposes intubated patients to VAP.

## Data Availability

All data produced in the present study are available upon reasonable request to the authors

## Figure Legends

**Supplementary Figure 1: Percentage of hospitalized patients with new positive SARS-CoV-2 PCR tests per month during 2020**. Based on the number of first-time positive SARS-CoV-2 PCR per month, relative to total patients tested in that month. Tests recorded in the data base began on March 10, 2020.

**Supplementary Figure 2: Rates of Respiratory Culture Ordering and Positivity in Intubated Patients**. A) Percentage of overall patients by COVID-19 status who are intubated. B) Percent of patients with respiratory cultures ordered who are intubated or non-intubated by COVID-19 status. C) Percent of patients with abnormal respiratory culture results who are intubated or non-intubated by COVID-19 status. D) Percentage of intubated, non-intubated or overall hospitalized patients who have respiratory cultures ordered by COVID-19 status. E) Percent of intubated, non-intubated or overall hospitalized patients with abnormal respiratory culture results out of the total number with cultures ordered by COVID-19 status. For intubated patients, only those respiratory cultures collected more than 48 hours after intubation were included. F) Percent of intubated or overall hospitalized patients with abnormal respiratory cultures out of total patients by COVID-19 status. For intubated patients, only those respiratory cultures collected more than 48 hours after intubation were included. * p < 0.05, ** p < 0.01, *** p < 0.001

**Supplementary Figure 3: Rates of antibiotic resistance by category**. Percentage of overall cultures with each type of resistance. Penicillin class includes penicillin-like beta lactams: penicillin, oxacillin and ampicillin; Cephalosporins include ceftriaxone and ceftazidime. * p < 0.05

